# PBPK modelling of dexamethasone in patients with COVID-19 and liver disease

**DOI:** 10.1101/2021.11.10.21266141

**Authors:** Maiara Camotti Montanha, Nicolas Cottura, Michael Booth, Daryl Hodge, Fazila Bunglawala, Hannah Kinvig, Sandra Grañana-Castillo, Andrew Lloyd, Saye Khoo, Marco Siccardi

## Abstract

The aim of the study was to apply Physiologically-Based Pharmacokinetic (PBPK) modelling to predict the effect of liver disease (LD) on the pharmacokinetics (PK) of dexamethasone (DEX) in the treatment of COVID-19. A whole-body PBPK model was created to simulate 100 adult individuals aged 18-60 years. Physiological changes (e.g., plasma protein concentration, liver size, CP450 expression, hepatic blood flow) and portal vein shunt were incorporated into the LD model. The changes were implemented by using the Child-Pugh (CP) classification system. DEX was qualified using clinical data in healthy adults for both oral (PO) and intravenous (IV) administrations and similarly propranolol (PRO) and midazolam (MDZ) were qualified with PO and IV clinical data in healthy and LD adults. The qualified model was subsequently used to simulate a 6 mg PO and 20 mg IV dose of DEX in patients with varying degrees of LD, with and without shunting. The PBPK model was successfully qualified across DEX, MDZ and PRO. In contrast to healthy adults, the simulated systemic clearance of DEX decreased (35% - 60%) and the plasma concentrations increased (170% - 400%) in patients with LD. Moreover, at higher doses of DEX, the AUC ratio between healthy/LD individuals remained comparable to lower doses. The exposure of DEX in different stages of LD was predicted through PBPK modelling, providing a rational framework to predict PK in complex clinical scenarios related to COVID-19. Model simulations suggest dose adjustments of DEX in LD patients are not necessary considering the low dose administered in the COVID-19 protocol.

## INTRODUCTION

Chronic liver disease (LD) is prevalent in 3 to 8% of patients suffering with COVID-19.^1^ Chronic LD has been associated with a higher rate of mortality in COVID-19 patients and can affect the drug distribution of several treatments.^2^ End stage chronic liver disease leads to cirrhosis which is characterised by the replacement of injured tissue with a collagenous scar and is accompanied by a loss of functional hepatocytes as well as a distortion in hepatic vasculature.^3,4^ The severity of liver disease can be classified using the CP score A, B, and C and is based on physiological and biological parameters.^5^ As the severity of liver disease increases, the distortion of the hepatic vasculature may lead to portal hypertension and in turn portacaval shunting. Shunting can significantly increase the bioavailability of a drug due to a decrease in first pass metabolism and this effect can be particularly relevant for drugs with a high first-pass extraction.^6^

DEX is a corticosteroid traditionally used in a wide range of conditions such as rheumatic or endocrine disorders for its anti-inflammatory and immunosuppressant effects,^7^ and can be used as a treatment for patients with severe COVID-19 disease.^8^ DEX has a relatively low hepatic extraction and is metabolised by the cytochrome P450 (CYP) enzymatic system, primarily by the CYP3A4 isoform, of which it is also a weak-moderate inducer.^7,9,10^ A previous study study showed reduced DEX clearance (CL) and prolonged half-life (t_1/2_) in individuals with LD compared with that in healthy subjects.^11^ However, DEX plasma concentrations, different forms of administration and detailed description of the individuals included in the study (e.g., LD severity according to CP score) are lacking.

PBPK modelling is a simulation approach with multiple applications and which is accepted by regulatory agencies primarily to evaluate enzyme-based drug-drug interactions (60% of submissions between 2008 and 2017).^12^ PBPK can account for changes in absorption, metabolism, distribution, and elimination (ADME), through the integration of *in vitro* data using *in vitro-in vivo* extrapolation (IVIVE) techniques for the prediction of PK in a cohort of virtual patients. The effect of liver disease on pharmacokinetics can be simulated considering a number of physiological changes as functional liver size, CYP450 expression, plasma protein binding and hepatic blood flow.^3,5^ The aim of this study was to use PBPK modelling to predict DEX PKs for the treatment of COVID-19 in patients with liver impairment.

## METHODS

A whole body PBPK model constructed using Simbiology v. 5.8.2, a product of MATLAB® R2019a v. 9.6.0 (MathWorks, Natick, MA, USA 2013), was used to generate a cohort of 100 individuals aged 18-60 years (50% female and 50% male). The following assumptions were made during simulations: (1) instant and uniform drug distribution (well-stirred model) across each compartment (tissue/organ); (2) no reabsorption of the drug from the colon; and (3) drug distribution was limited by blood flow. No ethical approval was required as results for this investigation were generated virtually.

### Anatomy and Physiology

The body mass index (BMI), body surface area (BSA), height and weight of the individuals were generated as described by de la Grandmaison *et al*.^13^ These values were used to allometrically calculate organs and tissues volumes through equations described by Bosgra *et al*.^14^ Density was used to calculate organs and tissues weights as described by Brown et al.^15^ Blood flows were calculated using percentage regional blood flows of the cardiac output described by the Environmental Protection Agency (EPA).^16^ To represent a LD population, changes to the physiological and biochemical parameters in the healthy adult model were made according to Johnson *et al*.^3^ The parameters and corresponding values are summarised in Table ***1***.

**Table 1.** Physiological and biochemical parameter changes in the liver disease model according to CP score (A, B, C). Johnson’s reported values.^3^

| Parameters | CP score |  |  |  |
| --- | --- | --- | --- | --- |
|  | Control | A | B | C |
| $Q_{pv}^*$ | 1.00 | 0.91 | 0.63 | 0.55 |
| $Q_{ha}^*$ | 1.00 | 1.40 | 1.62 | 1.91 |
| $Q_{cc}^*$ | 1.00 | 1.16 | 1.32 | 1.40 |
| $Q^*$ | 1.00 | 1.00 | 1.00 | 1.00 |
| $CL_{int,CYP3A4}^*$ | 1.00 | 0.59 | 0.39 | 0.25 |
| $CL_{int,CYP1A2}^*$ | 1.00 | 0.63 | 0.26 | 0.12 |
| $CL_{int,CYP2D6}^*$ | 1.00 | 0.76 | 0.33 | 0.11 |
| $CL_{int,CYP2C19}^*$ | 1.00 | 0.32 | 0.26 | 0.12 |
| $CL_{int,UGT}$ | 1.00 | 1.00 | 1.00 | 1.00 |
| $Ab_{CYP3A4,gut}^*$ | 1.00 | 0.84 | 0.57 | 0.35 |
| Albumin (g/L) | 44.70 | 41.10 | 33.90 | 26.30 |
| $W_{liver}^*$ | 1.00 | 0.81 | 0.65 | 0.53 |
| $GFR^*$ | 1.00 | 0.76 | 0.63 | 0.60 |

\*Values are a fraction of the control. $CL_{int}$ , intrinsic clearance of specific enzyme. $Ab_{CYP3A4,gut}$ , abundance of CYP3A4 in the intestinal tissue. $Q_{cc}$ , cardiac output. $Q_{ha}$ , blood flow of the hepatic artery. $Q_{pv}$ , blood flow of the portal vein. $Q$ , blood flow of other organs. $W_{liver}$ , weight of the liver.

### Oral absorption

Oral absorption was simulated using a compartmental absorption and transit model.^14^ The drug absorption rate constant (*K*_a_) was calculated using the effective permeability (P_eff_) based on the *in vivo* regional jejunal permeability in humans for PRO.^17^ For MDZ and DEX values observed in the literature for *K*_a_ were applied. The parameters are described in Table 2.

**Table 2.** Physiochemical and pharmacokinetic characteristics of dexamethasone, midazolam, and propranolol.

| Parameter | Dexamethasone | Midazolam | Propranolol |
| --- | --- | --- | --- |
| **Physiochemical** |  |  |  |
| Molecular weight (g/mol) | 392.46 <sup>7</sup> | 325.77 <sup>25</sup> | 259.34 <sup>49</sup> |
| $f_{up}$ | 0.23 <sup>7</sup> | 0.03 <sup>25</sup> | 0.1 <sup>49</sup> |
| $f_{ugut}$ | 1 | 1 | 1 |
| logP | 1.83 <sup>9</sup> | 3.89 <sup>25</sup> | 3.48 <sup>49</sup> |
| pKa (basic) | 12.42 <sup>7</sup> | 6.57 <sup>25</sup> | 9.5 <sup>49</sup> |
| R | 0.93 <sup>35</sup> | 0.55 <sup>29</sup> | 0.76 <sup>49</sup> |
| $K_a$ (h<sup>-1</sup>) | 1.9 <sup>9</sup> | 3.18 <sup>50</sup> | - |
| $P_{eff}$ (10<sup>-4</sup> cm/s) | - | - | 3.5 <sup>51</sup> |
| **Metabolism** |  |  |  |
| CL<sub>int,CYP3A4gut</sub> (μL/min/pmol) | 0.12 <sup>9</sup> | 1.7 <sup>18</sup> | - |
| CL<sub>int,CYP3A4liver</sub> (μL/min/pmol) | 0.12 <sup>9</sup> | 2.7 <sup>17</sup> | - |
| CL<sub>int,CYP1A2liver</sub> (μL/min/pmol) | - | - | 1.76 <sup>52</sup> |
| CL<sub>int,CYP2D6liver</sub> (μL/min/pmol) | - | - | 31.6 <sup>52</sup> |
| CL<sub>int,CYP2C19gut</sub> (μL/min/pmol) | - | - | 0.729 <sup>53</sup> |
| CL<sub>int,CYP2C19liver</sub> (μL/min/pmol) | - | - | 0.729 <sup>53</sup> |
| CL<sub>int,UGTliver</sub> (μL/min/mg) | - | - | 70.6 <sup>52</sup> |
| **CYP3A4 induction** |  |  |  |
| E<sub>max</sub> | 6.6 <sup>54</sup> | - | - |
| EC<sub>50</sub> (μM) | 51.22 <sup>54</sup> | - | - |
| **Distribution** |  |  |  |
| Vd correction factor | 0.6 | 0.2 | - |
| **Elimination** |  |  |  |
| CL<sub>renal</sub> (L/h) | 1.57 <sup>30</sup> |  |  |

$f_{up}$ , fraction of drug unbound in plasma. $f_{ugut}$ , fraction of drug unbound in the gut. logP, partition coefficient between water and octanol. pKa, acid dissociation constant. R, blood to plasma ratio. $K_a$ , absorption constant rate. $P_{eff}$ , effective permeability. CL<sub>int</sub>, intrinsic clearance. CYP, cytochrome P450. UGT, UDP-glucuronosyltransferases. EC<sub>50</sub> concentration of inducer producing 50 % of maximum induction, E<sub>max</sub> maximum induction. Vd, volume of distribution. CL<sub>renal</sub>, renal clearance.

### Intestinal metabolism

The clearance of MDZ, PRO and DEX in the gut (CLgut) were calculated considering the intrinsic clearance (Cl_int_) and abundance of the enzyme involved in the metabolism of each drug in the intestinal tissue (Eq. (1)):

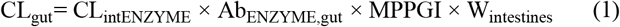

Where CL_intENZYME_ is the Cl_int_ of CYP3A4 for MDZ and DEX, and the Cl_int_ of CYP2C19 for PRO, Ab_ENZYME,gut_ is its relative abundance in the intestinal tissue (Ab_CYP3A4,gut_ = 19.2 pmol/mg,^18^ Ab_CYP2C19,gut_ = 2.1 ± 0.1 pmol/mg ^19^), MPPGI is the amount of microsomal protein per gram of intestine (MPPGI = 2.7 mg/g ^18^), and W_intestines_ is the weight of the intestines. The Cl_int_ of each enzyme is described in Table 2. CYP2D6 and CYP1A2 was not considered in the gut for PRO since its contribution to total intestinal CYP is minimal (<1%).^20^

The fraction of drug escaping gut metabolism and transitioning to the liver (F_g_) was computed with the following equation (Eq. (2)):

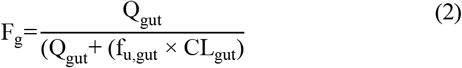

Where Q_gut_ represents the blood flow to the gut, and f_u,gut_ is the fraction unbound of the drug in the gut, considered equal 1 in the model.^21^

### Hepatic metabolism

Similarly, to the gut, the intrinsic clearance of each enzyme involved in the hepatic metabolism of MDZ, PRO and DEX were scaled up to the whole liver (CL_int,liver_) considering the equation below (Eq. (3)):

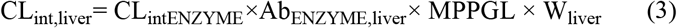

Where CL_intENZYME_ is the Cl_int_ of CYP3A4 for MDZ and DEX, and the Cl_int_ of CYP1A2, CYP2D6, CYP2C19 and UDP-glucuronosyltransferases (UGT) for PRO, Ab_ENZYME,liver_ is the abundance of the enzyme in the liver (Ab_CYP3A4,liver_ = 155 pmol/mg,^22^ Ab_CYP1A2,liver_ = 29.4 ± 29.6 pmol/mg,^23^ Ab_CYP2D6,liver_ = 11.9 ± 13.2 pmol/mg,^23^ Ab_CYP2C19,liver_ = 17.8 ± 3.3 pmol/mg ^24^) and MPPGL is the amount of microsomal protein per gram of liver, W_liver_ is the weight of the liver. The Cl_int_ of CYP3A4 and CYP2C19 was considered the same in the gut and liver for DEX and PRO, respectively, since no data specific to the gut were available. The intrinsic clearance of CYP3A4 in the gut and liver for MDZ were considered different All parameters are described in Table 2. For PRO an additional CL_int,liver_ was assumed to account for UGT metabolism (Eq. (4)):

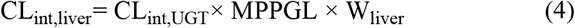

The MPPGL was calculated according to equation reported by Barter et al (Eq. (5)) ^25^:

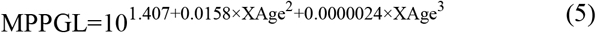

The total hepatic intrinsic clearance (∑CL_int,liver_) was considered as the sum of all enzymes involved in the metabolism. The hepatic systemic clearance (CL_hep_) was calculated considering blood flow and the total ∑CL_int,liver_ (Eq. (6)):

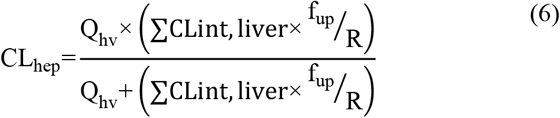

Where Q_hv_ is the hepatic blood flow rate, f_up_ is the fraction of drug unbound in plasma and R is the blood to plasma ratio. The fraction of drug that escapes hepatic metabolism and reaches the systemic circulation (F_h_) is represented by the following equation (Eq. (7)):

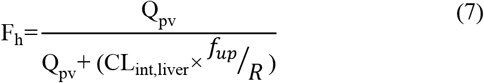

Where Q_pv_ is the blood flow rate of the portal vein.

Given DEX`s auto-induction of CYP3A4, induction of CYP in the intestine and liver were calculated from using the following equation (Eq. (8)):

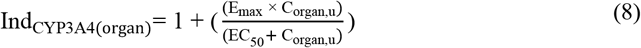

Where E_max_ is the maximum enzyme activity, C_organ,u_ is the average unbound drug concentration in the intestinal and liver tissues and EC_50_ is the DEX concentration required to reach half of the maximum enzyme activity. Then the CL_gut_ and CL_int,liver_ (equations 1 and 3) was multiplied by Ind_CYP3A4,organ_.

Portocaval shunting was incorporated into the LD model by implementing a shunt index that considers the varying levels of shunting associated with the different severities of liver disease as well as the serum total bile acid concentrations in the peripheral vein.^26,27^ The fraction of drug that bypasses the liver due to shunting (F_shunt_) is represented by the following (Eq. (9)):

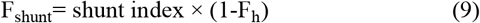

### Distribution

Drug distribution was calculated using first-order differential equations, with the volume of distribution (Vd) computed using the tissue to plasma ratio (TP) of each organ and the volume of each organ compartment.^28^ A correction factor (Table 2) was applied to the Vd of MDZ and DEX via curve-fitting method to match observed Vd values in the literature.^7,29^ The physiochemical properties of the drugs used in the models are detailed in Table 2.

### Elimination

Elimination of MDZ and PRO were considered as exclusively hepatic however, for DEX an additional renal clearance amounting to 10% of the systemic clearance was applied in accordance with the literature (Table 2).^30^ A liver impairment scaling factor for renal function was applied as described in Table 1.

### Model qualification

The model was firstly qualified for DEX, MDZ and PRO in a healthy population followed by qualification in a LD population for MDZ and PRO. The model qualification was extended to MDZ and PRO as the availability of observed clinical data across CP scores is incomplete and not all stages of LD and shunt index have been fully described. Specifically, MDZ was chosen due to its similarity in metabolic pathway to DEX, as both drugs are predominantly metabolised by CYP3A4. However, PK data for MDZ were not available for LD individuals with different CP scores, so a mixture of patients with different liver disease degrees was used.^31^ Therefore, to validate CP-A, -B and -C and the shunt index incorporated into the LD model individually, PRO was chosen due to the availability of observed clinical data reflecting these scenarios.^32^ A schematic representation of this workflow is shown in **Error! Reference source not found**.. The model qualification was performed according to recommendations of the European Medicine Agency (EMA) ^33^ and was considered validated when the mean of simulated PK parameters for each drug was less than two-fold of the observed clinical mean and the absolute average fold error (AFE) was below 2.^34^ The doses and regimens of the drugs were chosen to reflect the clinical studies used to validate the model.^31,32,35^ Due to the type of clinical data available for MDZ, simulated PK parameters for MDZ in the LD population were calculated considering the mean of conditions CP-A, -B and -C in order to reflect the clinical data sets. For PRO, clinical data were reported for 15 individuals alongside their respective CP score and shunt index thus, 100 simulations were carried out for each individual with their specific age, weight, CP score and shunt index implemented in the LD model.^32^ The mean PK parameters for PRO across all individuals were also calculated and compared for both simulated and observed clinical data.

### Predictions

The PBPK model was used to predict the PK of DEX in virtual populations with varying degrees of liver disease, classified according to CP scores (A, B and C). The varying levels of portacaval-shunting associated with liver disease considered in the simulations was an aleatory linearly spaced range with minimum value of 0.1 and maximum value of 0.7 as previously described.^26^

The dosages selected for the simulations were in line with current COVID-19 protocols stipulated by the National Institute for Health and Care Excellence; 6 mg dose once a day PO for 7 – 10 days; 6 mg dose once a day IV for 7 – 10 days.^36^ To further evaluate high doses of DEX, simulations with 20 mg were made. The PK parameters were calculated considering steady-state plasma concentration on 10^th^ day in accordance with the COVID-19 protocol.

## RESULTS

### Model qualification

The PBPK model was successfully qualified for all drugs in healthy (MDZ, PRO and DEX) and LD individuals (MDZ and PRO) according to the selected criteria. The simulated and observed PK parameters for each drug as well as the AFE values are presented in Table 3, as previously described. The plasma concentration-time profiles of each drug are described in the supplementary material (Figure S1-S5).

**Table 3.** Qualification of the PBPK model in healthy and liver disease individuals for midazolam, propranolol, and dexamethasone.

| Population | MDZ 7.5 mg IV |  |  |  | MDZ 15 mg oral |  |  |  |
| --- | --- | --- | --- | --- | --- | --- | --- | --- |
|  | Variables | Observed | Predicted | AFE | Variables | Observed | Predicted | AFE |
| Healthy | AUC<sub>0-inf</sub> (ng·h/mL) | 298<sup>31</sup> | 304.45 | 1.02 | AUC<sub>0-inf</sub> (ng·h/mL) | 362<sup>31</sup> | 260.50 | 1.39 |
|  | t<sub>1/2</sub> (h) | 3.8<sup>31</sup> | 3.32 | 1.15 | C<sub>max</sub> (ng/mL) | 62.83<sup>31</sup> | 54.66 | 1.15 |
|  | CL (mL/h/kg) | 337.8<sup>31</sup> | 328.46 | 1.03 | t<sub>max</sub> (h) | 0.75<sup>31</sup> | 0.75 | 1.00 |
| LD | AUC<sub>0-inf</sub> (ng·h/mL) | 543<sup>31</sup> | 468.39 | 1.16 | AUC<sub>0-inf</sub> (ng·h/mL) | 576<sup>31</sup> | 451.29 | 1.28 |
|  | t<sub>1/2</sub> (h) | 7.36<sup>31</sup> | 5.06 | 1.45 | C<sub>max</sub> (ng/mL) | 96.86<sup>31</sup> | 69.88 | 1.39 |
|  | CL (mL/h/kg) | 200.4<sup>31</sup> | 213.50 | 1.07 | t<sub>max</sub> (h) | 0.75<sup>31</sup> | 0.75 | 1.00 |
| PRO 1 mg IV |  |  |  |  | PRO 40 mg oral |  |  |  |
| Healthy | AUC<sub>0-inf</sub> (ng·min/mL) | 979<sup>32</sup> | 1166.01 | 1.19 | AUC<sub>0-inf</sub> (ng·min/mL) | 8930<sup>32</sup> | 10907.92 | 1.22 |
|  | t<sub>1/2</sub> (min) | 205<sup>32</sup> | 312.63 | 1.53 | C<sub>max</sub> (ng/mL) | 28<sup>32</sup> | 23.73 | 1.18 |
|  | CL (mL/min) | 1187<sup>32</sup> | 857.62 | 1.38 | t<sub>max</sub> (min) | 180<sup>32</sup> | 120.00 | 1.50 |
| LD | AUC<sub>0-inf</sub> (ng·min/mL) | 1778<sup>32</sup> | 1676 | 1.06 | AUC<sub>0-inf</sub> (ng·min/mL) | 47260<sup>32</sup> | 34258 | 1.38 |
|  | t<sub>1/2</sub> (min) | 641<sup>32</sup> | 398 | 1.61 | C<sub>max</sub> (ng/mL) | 65<sup>32</sup> | 60 | 1.08 |
|  | CL (mL/min) | 833<sup>32</sup> | 597 | 1.40 | t<sub>max</sub> (min) | 180<sup>32</sup> | 120 | 1.50 |
| DEXA 5 mg IV |  |  |  |  | DEXA 4.5 mg oral |  |  |  |
| Healthy | AUC<sub>0-inf</sub> (ng·h/mL) | 246<sup>35</sup> | 216.13 | 1.14 | AUC<sub>0-inf</sub> (ng·h/mL) | 239<sup>35</sup> | 238.15 | 1.00 |
|  | t<sub>1/2</sub> (h) | 4.1<sup>35</sup> | 3.58 | 1.15 | C<sub>max</sub> (ng/mL) | 38<sup>35</sup> | 34.64 | 1.09 |
|  | CL (mL/h/kg) | 243<sup>35</sup> | 330.49 | 1.36 | t<sub>max</sub> (h) | 2<sup>35</sup> | 2.00 | 1.10 |
|  | Vd (L/kg) | 1.4<sup>35</sup> | 1.71 | 1.22 | t<sub>1/2</sub> (h) | 4<sup>35</sup> | 3.59 | 1.11 |

Data are presented as the mean as described in section methods-model qualification. MDZ, midazolam. PRO, propranolol. DEXA, dexamethasone. $AUC_{0-\infty}$ , area under the plasma concentration-time curve over a dosing interval. $C_{\max}$ maximum plasma concentration. $t_{1/2}$ , half-life time. CL, clearance. $t_{\max}$ , time to maximum plasma concentration. Vd, volume of distribution. LD, liver dysfunction. AFE, average fold error.

### Predictions

The predicted PKs of DEX are shown in Table 4. The PBPK model for both the PO and IV administration of DEX predicted a decrease in the CL of DEX and increase in plasma concentration of DEX for patients with LD of all CP scores and shunt indexes. However, the exposure of DEX was found to be higher in patients with advanced CP scores, as shown in Figure 2. Furthermore, plasma concentrations of DEX were slightly higher in individuals with portal-systemic shunt compared to individuals with no shunting during PO administration. In Table 4, plasma concentrations are expressed as unbound in comparison to total plasma concentration shown in table 3. The shunting index had no effect when DEX was administered IV (**Error! Reference source not found**.) as the portal-systemic shunt primarily affects first-pass metabolism. When exploring higher doses of DEX it was found that the AUC ratio between healthy and LD individuals remained comparable to lower doses, as described in Table 4.

**Table 4.** Predictions of dexamethasone pharmacokinetics in virtual populations with varying degrees of liver disease.

| Parameters | Description | IV Dose (6 mg) |  |  | Oral Dose (6 mg) |  |  |
| --- | --- | --- | --- | --- | --- | --- | --- |
|  |  | CP-A | CP-B | CP-C | CP-A | CP-B | CP-C |
| AUC<sub>0-24</sub> | No Shunting | 141.37 (18) | 201.24 (15) | 315.77 (16) | 127 (20) | 180.20 (19) | 277.08 (16) |
|  | Ratio (No Shunting/Healthy) | 1.68 | 2.39 | 3.74 | 1.72 | 2.44 | 3.76 |
|  | Shunting | 144.24 (19) | 204.96 (17) | 307.63 (15) | 133.29 (18) | 194.81 (18) | 298.97 (17) |
|  | Ratio (Shunting/Healthy) | 1.71 | 2.43 | 3.65 | 1.81 | 2.64 | 4.06 |
| CL | No Shunting | 10.60 (19) | 8.34 (16) | 6.45 (17) | 11.80 (20) | 9.31 (19) | 7.35 (16) |
|  | Ratio (No Shunting/Healthy) | 0.65 | 0.51 | 0.39 | 0.63 | 0.50 | 0.39 |
|  | Shunting | 10.39 (20) | 8.19 (18) | 6.62 (15) | 11.25 (18) | 8.62 (18) | 6.82 (17) |
|  | Ratio (Shunting/Healthy) | 0.64 | 0.50 | 0.40 | 0.60 | 0.46 | 0.36 |
|  |  | IV Dose (20 mg) |  |  | Oral Dose (20 mg) |  |  |
| AUC<sub>0-24</sub> | No Shunting | 459.50 (17) | 669.14 (17) | 997.97 (17) | 419.20 (20) | 596.94 (20) | 894.69 (20) |
|  | Ratio (No Shunting/Healthy) | 1.68 | 2.44 | 3.65 | 1.73 | 2.47 | 3.70 |
| CL | No Shunting | 10.87 (18) | 8.36 (17) | 6.81 (18) | 11.92 (20) | 9.37 (20) | 7.59 (20) |
|  | Ratio (No Shunting/Healthy) | 0.65 | 0.50 | 0.41 | 0.63 | 0.49 | 0.40 |

Data are presented as the mean (coefficient of variation, %). AUC<sub>0-24</sub>, area under the plasma concentration-time curve over a dosing interval considering fraction of drug unbound to protein (ng·h /mL). CL, clearance (L/h). CP-A, CP-B and CP-C correspond to the Child-Pugh score.

**Figure 1.**
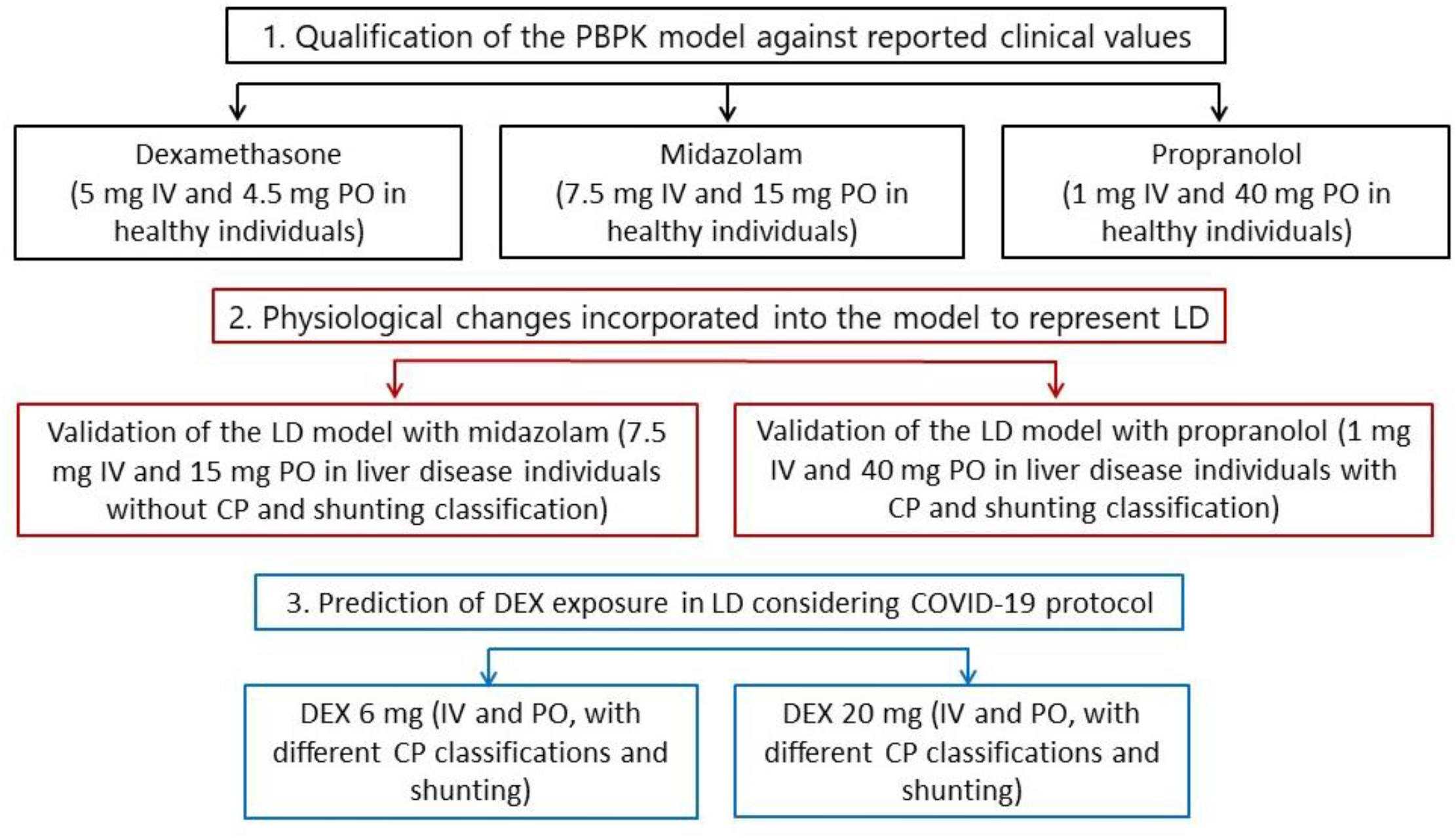
Overall step-by-step workflow representing the PBPK modelling qualification and predictions. IV, intravenous. PO, oral. LD, liver disease. CP, Child-Pugh. DEX, dexamethasone.

**Figure 2.**
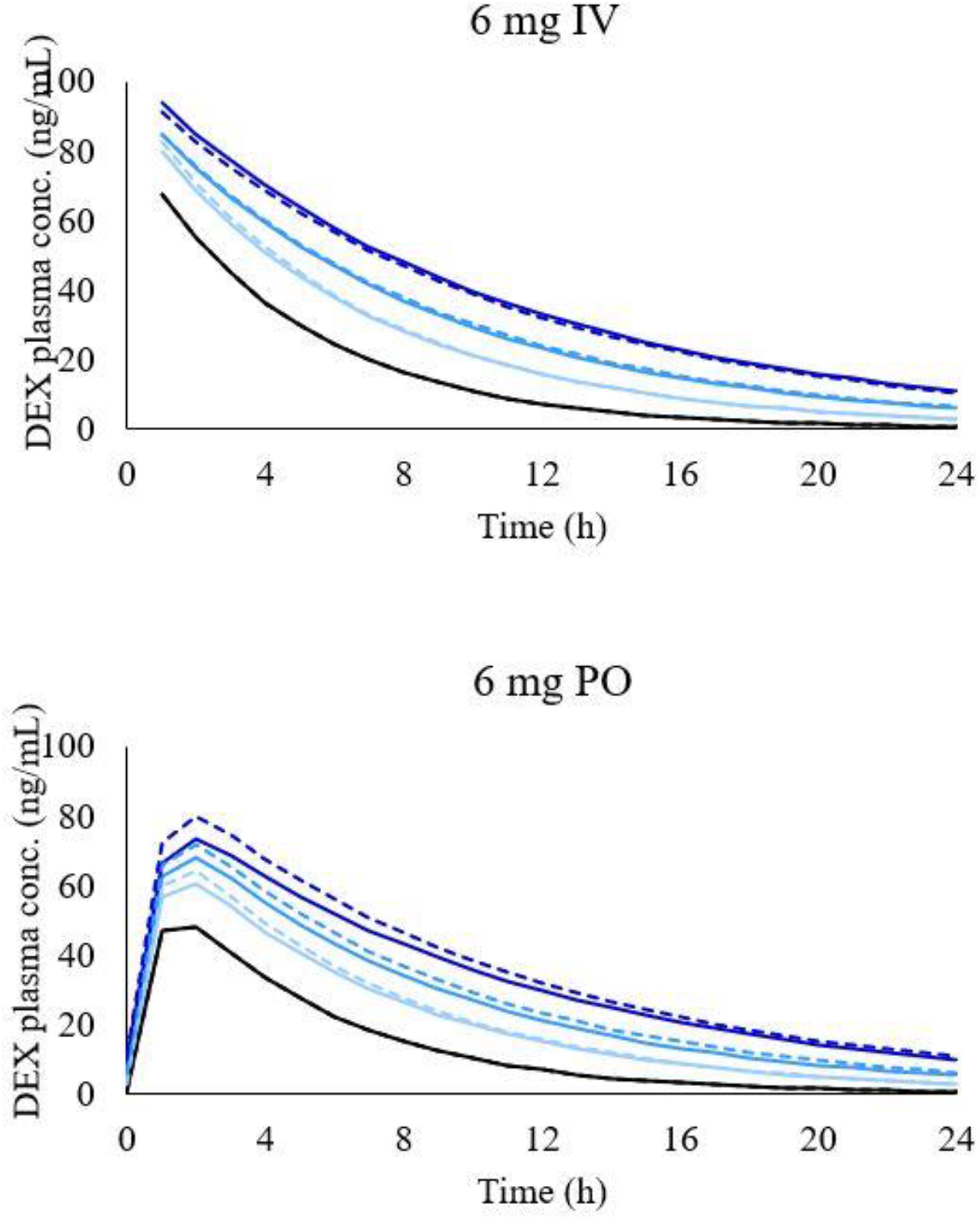
Simulated concentration-time profile of DEX in different LD conditions after 6 mg intravenous administration (graph on the top) and 6 mg oral administration (graph on the bottom). Black line, healthy individuals. Light blue line, CP-A condition, blue line, CP-B condition, and dark-blue line, CP-C condition. Dashed lines represent simulations with shunting and solid lines with no shunting.

## DISCUSSION

The clinical management of individuals with LD is challenging. The PK of DEX in LD patients has been partially described through a clinical study, showing a reduced clearance and increased half-life, but no information of total exposure (AUC0-inf) and PK profiles were available.^11^ The PBPK model described herein simulated DEX PK in different stages of LD with various grades of shunting, providing evidence-based guidance towards the clinical management of COVID-19 in LD patients.

Overall, the plasma concentration of DEX is expected to increase with liver impairment. The PBPK model was successfully validated and predicted an increase in AUC0-24 of 172% (181% with shunt), 244% (264% with shunt), and 376% (406% with shunt) compared to healthy individuals for CP-A, CP-B, and CP-C, respectively when DEX was administered orally. Furthermore, the corresponding clearance values were predicted to decrease approximately 35%, 50% and 60% in comparison to healthy individuals for CP-A, CP-B, and CP-C, respectively. The trend for CL to decrease was comparable to that previously found in the DEX clinical trial in LD patients.^11^ Furthermore, although first-pass metabolism can represent a relevant process in DEX PK, the predicted difference in AUC0-24 and CL between IV and PO administrations in LD individuals was minimal and is likely due to the high bioavailability (70-78%) of DEX.^7^ Additionally, first-pass metabolism is thought to be impacted by the shunt effect, increasing a drug’s bioavailability, yet the impact of shunting on the AUC0-24 and CL of DEX remained minimal. Simulations of DEX 20 mg IV and PO once a day were made since higher doses are being investigated to present clinical improvement and decrease in inflammatory biomarkers in patients hospitalized with COVID-19, but remain unproven.^37^ However, the AUC ratio between healthy and individuals with LD remained comparable to the lower dose (6 mg once a day), showing linear PK. For this reason, no simulation with shunt effect was performed with the higher dose since the same behaviour of lower dose is expected. According to the FDA and the EMA guidance for industry, PK studies should be conducted in patients with impaired hepatic function to evaluate whether a dose adjustment is necessary,^38,39^ yet the number of drugs that provide this specific recommendation for dosage adjustment based on different hepatic functions is very limited.^40^ This LD PBPK model could be applied to evaluate other drugs for use in COVID-19, such as anticoagulants, other corticosteroids, antiviral agents, antibiotics, anti-inflammatory drugs.^41^

According to the FDA guidelines, dosage adjustments should be recommended when a two-fold or greater increase in the AUC is observed.^38^ However, dose adjustment in COVID-19 patients is complex, defining a multifactorial scenario for which polypharmacy and pre-existing conditions should be considered alongside the risk of potential drug-drug interactions (DDIs) which may lead to altered PK. Moreover, the use of support resources in the care of COVID-19 patients, such as renal replacement, ventilation, volume replacement, can affect drug ADME generating further complexities in the assessment of dosing strategies.^42^

There are some aspects in the application of DEX for the treatment of COVID-19 that should be considered when analysing the data from this study. Firstly, severe COVID-19 is associated with a systemic hyper=-inflammation state with increased cytokine levels and highly elevated C-reactive protein (CRP) all of which are known to impact drug PK through the downregulation of CYP isoenzymes.^43-45^ The current PBPK model does not incorporate these mechanisms due to limited data/ability to verify the model. Secondly, DEX has been co-administered with tocilizumab for the treatment of COVID-19, however, the DDI between these drugs has not been studied in LD individuals. Tocilizumab has been reported to inhibit interleukin 6 (IL-6), increasing the activity of CYP450 enzymes and therefore producing increased metabolism of drugs that are CYP450 substrates.^46^ This pharmacodynamic DDI may compensate for the decreased CL observed in LD condition. Finally, corticosteroids such as DEX can present side effects relating to the central nervous system. In most cases the side effects occur within the 5 first days of treatment, however, the psychiatric symptoms tend to begin after 11 days and become more pronounced with extended periods of treatment. The psychiatric side effect also appears to be dose-dependent, occurring more often for doses up to 80 mg daily.^47^ Whilst side effects need to be monitored during the administration of DEX, no dose adjustments seem necessary in healthy or LD patients.

Though the PK of DEX in LD was successfully predicted, the model is characterized by some limitations. Although a direct and proportional relationship between CP score and increase in the plasma drug exposure was assume in the model, as demonstrated before by Johnson *et al*.,^3^ and also a linear correlation between portal vein shunt index and serum total bile acid concentrations in the peripheral vein,^27^ outliers individuals will not be represented by this assumption as demonstrated in supplementary table S2. Furthermore, the prevalence of spontaneous portosystemic shunt (SPS) increases as liver function injures, probably as an effect of damaging portal hypertension.^26^ However, large-SPS can be present in CP-A individuals as no SPS or small-SPS can be present in CP-C individuals.^26^ For this reason, the varying levels of portacaval-shunting associated with LD considered in the simulations was an aleatory linearly spaced range between 0.1 and 0.7. Other factors such as inter-individual variability, polymorphism, age (e.g., propranolol showed greater plasma level in elderly compared to young individuals),^48^ and unknown LD physiopathology mechanisms not represented in the model corroborate the challenge of qualifying the LD PBPK model against specific CP classifications.

## CONCLUSION

An increased exposure of DEX across varying stages of LD was predicted using PBPK modelling. Although DEX exposure was predicted to be more than 3 times higher in CP-C individuals, no dose adjustments seem necessary in patients with LD considering DEX’s low hepatic extraction, the low dose administered in the COVID-19 protocol and short period of treatment (10 days), and the therapeutic index of DEX. This study provides *in silico* evidence-based guidance towards the management of complex clinical scenarios related to COVID-19 and provides a rational framework for future PBPK modelling applications in LD patients. Further PBPK modelling initiatives would be necessary to evaluate the net effect of both LD and inflammatory physiological alterations on the PK of drugs used in the treatment of COVID-19.

### Study Highlights

- To propose a PBPK model capable of simulating the PK of drugs in LD patients classified according to the Child-Pugh system.
- To integrate portacaval-shunting associated with LD in a PBPK model.
- To predict DEX exposure in LD patients considering the dose administered in the COVID-19 protocol.

## Data Availability

All data produced in the present work are contained in the manuscript.

## Author contributions

All authors contributed to the overall concept of the model. M.C., N.C. and M.B. performed the modelling design, qualification, and application of the model. M.C., M.B. and M.S. wrote the manuscript with support from all the other authors. All authors reviewed and contributed to the final manuscript.

