## Supplemental material for "PBPK modelling of dexamethasone in patients with COVID-19 and liver disease"

**Supplementary material**

**Figure S1.** Simulated *versus* observed ^1^ mean plasma concentration of midazolam after administration of 7.5 mg intravenously (IV) and 15 mg orally (PO) in a healthy population.

Midazolam IV 7.5 mg - Healthy

MDZ plasma conc. (ng/mL)

| Time (h) | Mean predicted |
| --- | --- |
| 0.083 | 122.365 |
| 0.16 | 96.5066 |
| 0.25 | 84.3194 |
| 0.5 | 68.7548 |
| 0.75 | 60.3935 |
| 1 | 54.5678 |
| 1.5 | 46.4554 |
| 2 | 40.6346 |
| 3 | 32.0282 |
| 4 | 25.5909 |
| 6 | 16.5347 |
| 8 | 10.7697 |
| 12 | 4.6586 |
| 16 | 2.06409 |
| 24 | 0.432656 |

| Time (h) | Mean observed |
| --- | --- |
| 0.083 | 183.309 |
| 0.25 | 118.564 |
| 0.5 | 79.4079 |
| 0.75 | 59.0456 |
| 1 | 52.2641 |
| 2 | 32.6464 |
| 4 | 18.0503 |
| 6 | 11.8802 |
| 8 | 9.30796 |
| 12 | 4.03213 |
| 16 | 2.51864 |
| 24 | 1.90572 |

| Time (h) | ±SD | SD- |
| --- | --- | --- |
| 0.083 | 144.242 | 100.488 |
| 0.16 | 115.304 | 77.7087 |
| 0.25 | 101.064 | 67.5744 |
| 0.5 | 81.5824 | 55.9272 |
| 0.75 | 71.0338 | 49.7533 |
| 1 | 63.9189 | 45.2166 |
| 1.5 | 54.2144 | 38.6964 |
| 2 | 47.2148 | 34.0543 |
| 3 | 36.7886 | 27.2679 |
| 4 | 29.1819 | 22 |
| 6 | 19.1238 | 13.9455 |
| 8 | 12.988 | 8.55145 |
| 12 | 6.24127 | 3.07593 |
| 16 | 3.07322 | 1.05495 |
| 24 | 0.784176 | 0.0811369 |

Midazolam PO 15 mg - Healthy

MDZ plasma conc. (ng/mL)

| Time (h) | Mean predicted |
| --- | --- |
| 0.083 | 8.88502 |
| 0.16 | 20.4913 |
| 0.25 | 32.1301 |
| 0.5 | 50.3953 |
| 0.75 | 54.6593 |
| 1 | 52.8346 |
| 1.5 | 45.0622 |
| 2 | 38.3049 |
| 3 | 29.4074 |
| 4 | 23.4797 |
| 6 | 15.3585 |
| 8 | 10.1325 |
| 12 | 4.47309 |
| 16 | 2.01024 |
| 24 | 0.427775 |

| Time (h) | Mean observed |
| --- | --- |
| 0 | 0.0001 |
| 0.25 | 6.845 |
| 0.5 | 51.0219 |
| 0.75 | 62.8262 |
| 1 | 52.8734 |
| 2 | 38.759 |
| 4 | 24.7847 |
| 6 | 17.2761 |
| 8 | 12.0463 |
| 12 | 7.20711 |
| 16 | 5.12702 |
| 24 | 2.87867 |

| Time (h) | ±SD | SD- |
| --- | --- | --- |
| 0.083 | 10.3571 | 7.41293 |
| 0.16 | 23.7341 | 17.2485 |
| 0.25 | 37.2566 | 27.0037 |
| 0.5 | 58.7865 | 42.0041 |
| 0.75 | 63.88 | 45.4386 |
| 1 | 61.6389 | 44.0304 |
| 1.5 | 52.2474 | 37.877 |
| 2 | 44.2967 | 32.313 |
| 3 | 34.1452 | 24.6697 |
| 4 | 27.5178 | 19.4417 |
| 6 | 18.5529 | 12.1641 |
| 8 | 12.7418 | 7.52325 |
| 12 | 6.15943 | 2.78674 |
| 16 | 3.04058 | 0.979907 |
| 24 | 0.782936 | 0.072614 |

**Figure S2.** Simulated *versus* observed ^1^ mean plasma concentration of midazolam after administration of 7.5 mg intravenously (IV) and 15 mg orally (PO) considering the mean of Child-Pugh (CP)-A, CP-B and CP-C liver disease classification.

Midazolam IV 7.5 mg (LD mean CP-A, B, C)

MDZ plasma conc. (ng/mL)

| Time (h) | Mean predicted |
| --- | --- |
| 0.083 | 130.527 |
| 0.16 | 105.365 |
| 0.25 | 93.1318 |
| 0.5 | 77.0559 |
| 0.75 | 68.3783 |
| 1 | 62.6539 |
| 1.5 | 55.2083 |
| 2 | 50.0945 |
| 3 | 42.467 |
| 4 | 36.4477 |
| 6 | 27.1438 |
| 8 | 20.3863 |
| 12 | 11.7481 |
| 16 | 6.94763 |
| 24 | 2.59866 |

| Time (h) | Mean observed |
| --- | --- |
| 0.083 | 159.452 |
| 0.25 | 96.1888 |
| 0.5 | 79.4079 |
| 0.75 | 66.7071 |
| 1 | 51.3611 |
| 2 | 38.1907 |
| 4 | 27.4248 |
| 6 | 19.6938 |
| 8 | 18.3676 |
| 12 | 11.4732 |
| 16 | 7.55135 |
| 24 | 5.32894 |

| Time (h) | ±SD | SD- |
| --- | --- | --- |
| 0.083 | 155.668 | 105.386 |
| 0.16 | 127.013 | 83.7159 |
| 0.25 | 111.878 | 74.3853 |
| 0.5 | 91.0235 | 63.0883 |
| 0.75 | 80.1978 | 56.5588 |
| 1 | 73.3615 | 51.9463 |
| 1.5 | 64.5852 | 45.8315 |
| 2 | 58.3991 | 41.7899 |
| 3 | 48.9334 | 36.0005 |
| 4 | 41.6088 | 31.2867 |
| 6 | 31.0816 | 23.206 |
| 8 | 23.9744 | 16.7981 |
| 12 | 14.9415 | 8.55469 |
| 16 | 9.58127 | 4.314 |
| 24 | 4.14562 | 1.05171 |

Midazolam PO 15 mg (LD mean CP-A, B, C)

MDZ plasma conc. (ng/mL)

| Time (h) | Mean predicted |
| --- | --- |
| 0 | 0 |
| 0.083 | 11.3836 |
| 0.16 | 25.4152 |
| 0.25 | 39.7021 |
| 0.5 | 63.328 |
| 0.75 | 69.8816 |
| 1 | 68.4471 |
| 1.5 | 59.7757 |
| 2 | 52.2515 |
| 3 | 42.8233 |
| 4 | 36.5687 |
| 6 | 27.3437 |
| 8 | 20.6512 |
| 12 | 12.0224 |
| 16 | 7.17612 |
| 24 | 2.73566 |

| Time (h) | Mean observed |
| --- | --- |
| 0 | 0.0001 |
| 0.25 | 48.505 |
| 0.5 | 81.7649 |
| 0.75 | 92.1978 |
| 1 | 91.3251 |
| 2 | 71.0497 |
| 4 | 46.7958 |
| 6 | 31.7254 |
| 8 | 21.6799 |
| 12 | 10.3479 |
| 16 | 5.07293 |
| 24 | 1.30632 |

| Time (h) | ±SD | SD- |
| --- | --- | --- |
| 0 | 0 | 0 |
| 0.083 | 13.1785 | 9.58857 |
| 0.16 | 29.3853 | 21.4451 |
| 0.25 | 46.0747 | 33.3295 |
| 0.5 | 73.9531 | 52.7029 |
| 0.75 | 81.486 | 58.2772 |
| 1 | 79.43 | 57.4643 |
| 1.5 | 68.8438 | 50.7075 |
| 2 | 60.1535 | 44.3496 |
| 3 | 49.5871 | 36.0595 |
| 4 | 42.6726 | 30.4647 |
| 6 | 32.7335 | 21.9539 |
| 8 | 25.5996 | 15.7029 |
| 12 | 16.1004 | 7.94447 |
| 16 | 10.3688 | 3.98343 |
| 24 | 4.54886 | 0.922465 |

**Figure S3.** Simulated *versus* observed ^2^ mean plasma concentration of propranolol after administration of 1 mg intravenously (IV) and 40 mg orally (PO) in a healthy population.

Propranolol IV 1 mg - Healthy

PRO plasma conc. (ng/mL)

| Time (h) | Mean predicted |
| --- | --- |
| 0.08 | 14.6665 |
| 0.25 | 4.13202 |
| 0.5 | 2.87376 |
| 1 | 2.2792 |
| 1.5 | 2.00361 |
| 2 | 1.81327 |
| 3 | 1.52924 |
| 4 | 1.31001 |
| 5 | 1.13072 |
| 6 | 0.980294 |
| 7 | 0.852207 |
| 8 | 0.742153 |
| 9 | 0.647069 |
| 10 | 0.56463 |
| 11 | 0.492995 |
| 12 | 0.430655 |
| 13 | 0.376351 |
| 14 | 0.329012 |
| 15 | 0.287721 |
| 16 | 0.251689 |
| 17 | 0.220235 |
| 18 | 0.192767 |
| 19 | 0.168773 |
| 20 | 0.147806 |
| 21 | 0.12948 |
| 22 | 0.113457 |
| 23 | 0.0994434 |
| 24 | 0.0871845 |
| 25 | 0.0764574 |
| 26 | 0.0670682 |
| 27 | 0.0588478 |
| 28 | 0.0516487 |
| 29 | 0.0453424 |
| 30 | 0.0398166 |
| 31 | 0.0349735 |
| 32 | 0.0307276 |
| 33 | 0.0270043 |
| 34 | 0.0237384 |
| 35 | 0.0208731 |
| 36 | 0.0183584 |
| 37 | 0.0161509 |
| 38 | 0.0142126 |
| 39 | 0.0125101 |
| 40 | 0.0110145 |
| 41 | 0.00970017 |
| 42 | 0.00854496 |
| 43 | 0.00752927 |
| 44 | 0.00663603 |
| 45 | 0.00585028 |
| 46 | 0.0051589 |
| 47 | 0.00455041 |
| 48 | 0.00401466 |

| Time (h) | Mean observed |
| --- | --- |
| 0.083 | 8.09953 |
| 0.25 | 5.24417 |
| 0.5 | 4.60964 |
| 1 | 2.96734 |
| 1.5 | 2.55676 |
| 2 | 2.16485 |
| 3 | 1.41835 |
| 5 | 1.1944 |
| 8 | 0.335925 |
| 14 | 0.186625 |
| 24 | 0.0186625 |
| 48 | 0.0559876 |

| Time (h) | ±SD | SD- |
| --- | --- | --- |
| 0.08 | 17.6251 | 11.708 |
| 0.25 | 5.06967 | 3.19437 |
| 0.5 | 3.57804 | 2.16949 |
| 1 | 2.82041 | 1.73799 |
| 1.5 | 2.44632 | 1.5609 |
| 2 | 2.18281 | 1.44372 |
| 3 | 1.80161 | 1.25687 |
| 4 | 1.52459 | 1.09543 |
| 5 | 1.30823 | 0.953206 |
| 6 | 1.13214 | 0.828447 |
| 7 | 0.985194 | 0.719219 |
| 8 | 0.860661 | 0.623646 |
| 9 | 0.754049 | 0.540088 |
| 10 | 0.662129 | 0.46713 |
| 11 | 0.582454 | 0.403535 |
| 12 | 0.513106 | 0.348204 |
| 13 | 0.452552 | 0.30015 |
| 14 | 0.399539 | 0.258485 |
| 15 | 0.353029 | 0.222414 |
| 16 | 0.312155 | 0.191224 |
| 17 | 0.276185 | 0.164286 |
| 18 | 0.244494 | 0.141041 |
| 19 | 0.216545 | 0.121001 |
| 20 | 0.191876 | 0.103737 |
| 21 | 0.170087 | 0.0888734 |
| 22 | 0.150829 | 0.0760843 |
| 23 | 0.1338 | 0.0650866 |
| 24 | 0.118734 | 0.0556348 |
| 25 | 0.105399 | 0.047516 |
| 26 | 0.0935906 | 0.0405458 |
| 27 | 0.0831307 | 0.034565 |
| 28 | 0.0738616 | 0.0294358 |
| 29 | 0.0656451 | 0.0250397 |
| 30 | 0.0583592 | 0.0212741 |
| 31 | 0.0518963 | 0.0180506 |
| 32 | 0.046162 | 0.0152932 |
| 33 | 0.0410725 | 0.0129361 |
| 34 | 0.0365541 | 0.0109228 |
| 35 | 0.0325415 | 0.0092046 |
| 36 | 0.0289772 | 0.0077396 |
| 37 | 0.02581 | 0.00649173 |
| 38 | 0.0229952 | 0.00542994 |
| 39 | 0.0204926 | 0.00452757 |
| 40 | 0.0182672 | 0.00376171 |
| 41 | 0.0162877 | 0.00311266 |
| 42 | 0.0145264 | 0.00256349 |
| 43 | 0.0129589 | 0.00209961 |
| 44 | 0.0115635 | 0.00170855 |
| 45 | 0.010321 | 0.00137958 |
| 46 | 0.0092143 | 0.00110351 |
| 47 | 0.00822835 | 0.000872464 |
| 48 | 0.00734967 | 0.00067965 |

Propranolol PO 40 mg - Healthy

PRO plasma conc. (ng/mL)

| Time (h) | Mean predicted |
| --- | --- |
| 0 | 0 |
| 0.08 | 0 |
| 0.25 | 0 |
| 0.5 | 0.962004 |
| 1 | 13.4239 |
| 1.5 | 23.7275 |
| 2 | 23.7302 |
| 3 | 18.9165 |
| 4 | 15.4807 |
| 5 | 13.0906 |
| 6 | 11.2246 |
| 7 | 9.68238 |
| 8 | 8.37821 |
| 9 | 7.26328 |
| 10 | 6.30436 |
| 11 | 5.47666 |
| 12 | 4.76065 |
| 13 | 4.14036 |
| 14 | 3.60246 |
| 15 | 3.13568 |
| 16 | 2.73038 |
| 17 | 2.37829 |
| 18 | 2.0723 |
| 19 | 1.80627 |
| 20 | 1.5749 |
| 21 | 1.37359 |
| 22 | 1.1984 |
| 23 | 1.04587 |
| 24 | 0.913029 |
| 25 | 0.797304 |
| 26 | 0.696453 |
| 27 | 0.608539 |
| 28 | 0.531877 |
| 29 | 0.465007 |
| 30 | 0.406661 |
| 31 | 0.355736 |
| 32 | 0.311276 |
| 33 | 0.272448 |
| 34 | 0.23853 |
| 35 | 0.20889 |
| 36 | 0.182983 |
| 37 | 0.160332 |
| 38 | 0.140522 |
| 39 | 0.123192 |
| 40 | 0.108027 |
| 41 | 0.0947537 |
| 42 | 0.0831322 |
| 43 | 0.0729545 |
| 44 | 0.0640389 |
| 45 | 0.0562266 |
| 46 | 0.0493794 |
| 47 | 0.0433767 |
| 48 | 0.038113 |

| Time (h) | Mean observed |
| --- | --- |
| 0.5 | 3.04821 |
| 1 | 16.5474 |
| 1.5 | 22.535 |
| 2 | 25.8009 |
| 3 | 21.8818 |
| 5 | 12.9549 |
| 8 | 6.20529 |
| 14 | 3.26594 |
| 24 | 0.870918 |
| 48 | 0.653188 |

| Time (h) | ±SD | SD- |
| --- | --- | --- |
| 0 | 0 | 0 |
| 0.08 | 0 | 0 |
| 0.25 | 0 | 0 |
| 0.5 | 1.28779 | 0.636223 |
| 1 | 17.3387 | 9.50921 |
| 1.5 | 30.5015 | 16.9535 |
| 2 | 30.4955 | 16.9649 |
| 3 | 24.7055 | 13.1274 |
| 4 | 20.3545 | 10.6068 |
| 5 | 17.248 | 8.9331 |
| 6 | 14.8085 | 7.64075 |
| 7 | 12.7941 | 6.57065 |
| 8 | 11.095 | 5.66138 |
| 9 | 9.64658 | 4.87997 |
| 10 | 8.40389 | 4.20483 |
| 11 | 7.33316 | 3.62016 |
| 12 | 6.40771 | 3.11359 |
| 13 | 5.60582 | 2.67489 |
| 14 | 4.90956 | 2.29537 |
| 15 | 4.30393 | 1.96743 |
| 16 | 3.7763 | 1.68446 |
| 17 | 3.31598 | 1.4406 |
| 18 | 2.91388 | 1.23072 |
| 19 | 2.56223 | 1.05031 |
| 20 | 2.25439 | 0.895402 |
| 21 | 1.98464 | 0.762545 |
| 22 | 1.74808 | 0.648713 |
| 23 | 1.54046 | 0.551278 |
| 24 | 1.3581 | 0.467955 |
| 25 | 1.19784 | 0.396765 |
| 26 | 1.05691 | 0.335996 |
| 27 | 0.932911 | 0.284167 |
| 28 | 0.823752 | 0.240002 |
| 29 | 0.727613 | 0.202402 |
| 30 | 0.642902 | 0.17042 |
| 31 | 0.568231 | 0.143242 |
| 32 | 0.502384 | 0.120169 |
| 33 | 0.444295 | 0.100602 |
| 34 | 0.393033 | 0.0840256 |
| 35 | 0.347781 | 0.0699995 |
| 36 | 0.30782 | 0.0581462 |
| 37 | 0.272522 | 0.0481428 |
| 38 | 0.241331 | 0.0397129 |
| 39 | 0.213764 | 0.0326208 |
| 40 | 0.189391 | 0.0266643 |
| 41 | 0.167836 | 0.0216715 |
| 42 | 0.148769 | 0.0174957 |
| 43 | 0.131897 | 0.0140116 |
| 44 | 0.116965 | 0.0111125 |
| 45 | 0.103746 | 0.00870711 |
| 46 | 0.0920406 | 0.00671827 |
| 47 | 0.0816732 | 0.00508026 |
| 48 | 0.072489 | 0.00373696 |

Propranolol IV 1 mg (Mean CP-A,B,C)

PRO plasma conc. (ng/mL)

| Time (h) | Mean predicted |
| --- | --- |
| 0.08 | 14.9055 |
| 0.25 | 4.74802 |
| 0.5 | 3.51417 |
| 1 | 2.91437 |
| 1.5 | 2.61832 |
| 2 | 2.39974 |
| 3 | 2.06201 |
| 4 | 1.79905 |
| 5 | 1.58256 |
| 6 | 1.39882 |
| 7 | 1.24008 |
| 8 | 1.10153 |
| 9 | 0.979868 |
| 10 | 0.872623 |
| 11 | 0.777859 |
| 12 | 0.69398 |
| 13 | 0.619639 |
| 14 | 0.553681 |
| 15 | 0.495107 |
| 16 | 0.443046 |
| 17 | 0.396737 |
| 18 | 0.355513 |
| 19 | 0.318787 |
| 20 | 0.286046 |
| 21 | 0.256835 |
| 22 | 0.230757 |
| 23 | 0.207458 |
| 24 | 0.186629 |
| 25 | 0.167994 |
| 26 | 0.151312 |
| 27 | 0.136367 |
| 28 | 0.122971 |
| 29 | 0.110955 |
| 30 | 0.10017 |
| 31 | 0.0904837 |
| 32 | 0.081779 |
| 33 | 0.0739515 |
| 34 | 0.0669087 |
| 35 | 0.0605682 |
| 36 | 0.0548565 |
| 37 | 0.0497084 |
| 38 | 0.0450657 |
| 39 | 0.0408764 |
| 40 | 0.0370942 |
| 41 | 0.0336777 |
| 42 | 0.0305898 |
| 43 | 0.0277976 |
| 44 | 0.0252714 |
| 45 | 0.0229847 |
| 46 | 0.0209138 |
| 47 | 0.0190374 |
| 48 | 0.0173364 |

| Time (h) | Mean observed |
| --- | --- |
| 0.083 | 8.09953 |
| 0.25 | 5.24417 |
| 0.5 | 4.60964 |
| 1 | 2.96734 |
| 1.5 | 2.55676 |
| 2 | 2.16485 |
| 3 | 1.41835 |
| 5 | 1.1944 |
| 8 | 0.335925 |
| 14 | 0.186625 |
| 24 | 0.0186625 |
| 48 | 0.0559876 |

| Time (h) | ±SD | SD- |
| --- | --- | --- |
| 0.08 | 15.2959 | 14.5151 |
| 0.25 | 5.04642 | 4.44961 |
| 0.5 | 3.82477 | 3.20357 |
| 1 | 3.22479 | 2.60396 |
| 1.5 | 2.93368 | 2.30295 |
| 2 | 2.71815 | 2.08133 |
| 3 | 2.38489 | 1.73912 |
| 4 | 2.1243 | 1.47379 |
| 5 | 1.90716 | 1.25797 |
| 6 | 1.71968 | 1.07796 |
| 7 | 1.55452 | 0.925644 |
| 8 | 1.4074 | 0.795673 |
| 9 | 1.2755 | 0.684238 |
| 10 | 1.15681 | 0.588432 |
| 11 | 1.04978 | 0.505934 |
| 12 | 0.953132 | 0.434828 |
| 13 | 0.865765 | 0.373512 |
| 14 | 0.786737 | 0.320625 |
| 15 | 0.71521 | 0.275004 |
| 16 | 0.650441 | 0.235652 |
| 17 | 0.591763 | 0.201711 |
| 18 | 0.538582 | 0.172443 |
| 19 | 0.490363 | 0.147211 |
| 20 | 0.446625 | 0.125466 |
| 21 | 0.406936 | 0.106734 |
| 22 | 0.370908 | 0.0906059 |
| 23 | 0.338189 | 0.0767271 |
| 24 | 0.308465 | 0.0647922 |
| 25 | 0.281451 | 0.0545368 |
| 26 | 0.256891 | 0.0457323 |
| 27 | 0.234553 | 0.0381813 |
| 28 | 0.21423 | 0.0317128 |
| 29 | 0.195731 | 0.0261789 |
| 30 | 0.178888 | 0.0214518 |
| 31 | 0.163547 | 0.0174207 |
| 32 | 0.149568 | 0.0139899 |
| 33 | 0.136827 | 0.0110764 |
| 34 | 0.125209 | 0.00860867 |
| 35 | 0.114612 | 0.00652453 |
| 36 | 0.104943 | 0.00477038 |
| 37 | 0.0961171 | 0.00329976 |
| 38 | 0.0880589 | 0.00207254 |
| 39 | 0.0806989 | 0.00105395 |
| 40 | 0.0739745 | 0.00021398 |
| 41 | 0.0678287 | -0.000473343 |
| 42 | 0.0622101 | -0.00103042 |
| 43 | 0.0570718 | -0.00147661 |
| 44 | 0.0523713 | -0.00182857 |
| 45 | 0.0480701 | -0.00210069 |
| 46 | 0.044133 | -0.00230536 |
| 47 | 0.0405281 | -0.00245325 |
| 48 | 0.0372265 | -0.00255364 |

**Figure S4.** Simulated *versus* observed ^2^ mean plasma concentration of propranolol after administration of 1 mg intravenously (IV) and 40 mg orally (PO) considering the mean of Child-Pugh (CP)-A, CP-B and CP-C liver disease classification.

Propranolol PO 40 mg (Mean CP-A,B,C)

PRO plasma conc. (ng/mL)

| Time (h) | Mean predicted |
| --- | --- |
| 0.08 | 0 |
| 0.25 | 0 |
| 0.5 | 4.86444 |
| 1 | 35.2191 |
| 1.5 | 60.4401 |
| 2 | 60.3897 |
| 3 | 50.6274 |
| 4 | 42.9314 |
| 5 | 37.3122 |
| 6 | 32.8557 |
| 7 | 29.1263 |
| 8 | 25.9187 |
| 9 | 23.1192 |
| 10 | 20.6554 |
| 11 | 18.4763 |
| 12 | 16.5432 |
| 13 | 14.8249 |
| 14 | 13.2951 |
| 15 | 11.9318 |
| 16 | 10.7155 |
| 17 | 9.62968 |
| 18 | 8.65944 |
| 19 | 7.79187 |
| 20 | 7.01557 |
| 21 | 6.32046 |
| 22 | 5.69763 |
| 23 | 5.1392 |
| 24 | 4.63818 |
| 25 | 4.18838 |
| 26 | 3.78432 |
| 27 | 3.42113 |
| 28 | 3.09446 |
| 29 | 2.80047 |
| 30 | 2.53574 |
| 31 | 2.29721 |
| 32 | 2.08217 |
| 33 | 1.88819 |
| 34 | 1.71313 |
| 35 | 1.55503 |
| 36 | 1.4122 |
| 37 | 1.28307 |
| 38 | 1.16629 |
| 39 | 1.0606 |
| 40 | 0.964922 |
| 41 | 0.878251 |
| 42 | 0.799704 |
| 43 | 0.728486 |
| 44 | 0.663882 |
| 45 | 0.605251 |
| 46 | 0.552017 |
| 47 | 0.503661 |
| 48 | 0.459717 |

| Time (h) | Mean observed |
| --- | --- |
| 0.5 | 9.03577 |
| 1 | 21.7729 |
| 1.5 | 40.9331 |
| 2 | 44.9611 |
| 3 | 55.9565 |
| 5 | 39.3002 |
| 8 | 31.5708 |
| 14 | 25.5832 |
| 24 | 11.4308 |
| 48 | 4.57232 |

| Time (h) | ±SD | SD- |
| --- | --- | --- |
| 0.08 | 0 | 0 |
| 0.25 | 0 | 0 |
| 0.5 | 7.20223 | 2.52665 |
| 1 | 47.8721 | 22.5662 |
| 1.5 | 81.8516 | 39.0287 |
| 2 | 81.9841 | 38.7953 |
| 3 | 69.9409 | 31.3139 |
| 4 | 60.3457 | 25.5171 |
| 5 | 53.3045 | 21.32 |
| 6 | 47.6809 | 18.0306 |
| 7 | 42.9249 | 15.3277 |
| 8 | 38.7809 | 13.0564 |
| 9 | 35.1119 | 11.1265 |
| 10 | 31.8339 | 9.47684 |
| 11 | 28.8901 | 8.0626 |
| 12 | 26.238 | 6.84848 |
| 13 | 23.8441 | 5.80567 |
| 14 | 21.6802 | 4.91005 |
| 15 | 19.7223 | 4.14118 |
| 16 | 17.9495 | 3.48156 |
| 17 | 16.3432 | 2.91613 |
| 18 | 14.8869 | 2.43194 |
| 19 | 13.5659 | 2.01781 |
| 20 | 12.3671 | 1.66407 |
| 21 | 11.2785 | 1.36238 |
| 22 | 10.2897 | 1.10554 |
| 23 | 9.3911 | 0.887293 |
| 24 | 8.57408 | 0.702272 |
| 25 | 7.83095 | 0.545818 |
| 26 | 7.15473 | 0.413912 |
| 27 | 6.53917 | 0.30308 |
| 28 | 5.97859 | 0.210323 |
| 29 | 5.46789 | 0.133051 |
| 30 | 5.00244 | 0.0690288 |
| 31 | 4.57809 | 0.0163282 |
| 32 | 4.19105 | -0.0267145 |
| 33 | 3.83792 | -0.0615338 |
| 34 | 3.51562 | -0.0893654 |
| 35 | 3.22134 | -0.111273 |
| 36 | 2.95257 | -0.128173 |
| 37 | 2.707 | -0.140852 |
| 38 | 2.48256 | -0.149986 |
| 39 | 2.27737 | -0.156156 |
| 40 | 2.0897 | -0.159859 |
| 41 | 1.91802 | -0.161521 |
| 42 | 1.76091 | -0.161506 |
| 43 | 1.6171 | -0.160124 |
| 44 | 1.4854 | -0.15764 |
| 45 | 1.36478 | -0.154279 |
| 46 | 1.25426 | -0.150231 |
| 47 | 1.15298 | -0.145657 |
| 48 | 1.06013 | -0.140693 |

**Figure S5.** Simulated *versus* observed ^3^ mean plasma concentration of dexamethasone after administration of 5 mg intravenously (IV) and 4.5 mg orally (PO) in a healthy population.

Dexamethasone IV 5 mg - Healthy

DEX plasma conc. (ng/mL)

| Time (h) | Mean predicted | Mean observed | ±SD | SD- |
| --- | --- | --- | --- | --- |
| 1 | 42.2695 | 37.813 | 50.3815 | 34.1575 |
| 2 | 34.3131 | 31.1419 | 39.7237 | 28.9026 |
| 3 | 27.9489 | 26.8178 | 31.682 | 24.2157 |
| 4 | 22.8 | 21.4098 | 25.5341 | 20.0658 |
| 6 | 15.2363 | 14.3862 | 17.1784 | 13.2942 |
| 8 | 10.2342 | 9.70869 | 11.9325 | 8.53591 |
| 10 | 6.90679 | 6.65618 | 8.3929 | 5.42068 |
| 12 | 4.68148 | 4.86632 | 5.93076 | 3.4322 |
| 23 | 0.588062 | 0.801819 | 0.905468 | 0.270656 |

Dexamethasone OD 4.5 mg - Healthy

DEX plasma conc. (ng/mL)

| Time (h) | Mean predicted | Mean observed | ±SD | SD- |
| --- | --- | --- | --- | --- |
| 0 | 0 | 0.001 | 0 | 0 |
| 1 | 33.7125 | 32.5806 | 39.5761 | 27.8489 |
| 2 | 34.6419 | 30.9677 | 40.5033 | 28.7805 |
| 3 | 29.236 | 29.5341 | 33.8849 | 24.587 |
| 4 | 24.0152 | 23.4409 | 27.6739 | 20.3564 |
| 6 | 16.1334 | 17.1685 | 18.6415 | 13.6253 |
| 8 | 10.8691 | 11.4337 | 12.7952 | 8.94293 |
| 10 | 7.34455 | 8.02867 | 8.88987 | 5.79923 |
| 12 | 4.97728 | 5.3405 | 6.21998 | 3.73459 |
| 23 | 0.613849 | 0.860215 | 0.919014 | 0.308684 |

**Table S1.** Mean simulated PK parameters of midazolam (MDZ) in different liver disease conditions.

|  | MDZ 7.5 mg IV | | | | | | MDZ 15 mg PO | | | | | |
| --- | --- | --- | --- | --- | --- | --- | --- | --- | --- | --- | --- | --- |
|  | AUC_0-inf_ | AFE | CL | AFE | t_1/2_ | AFE | AUC_0-inf_ | AFE | C_max_ | AFE | t_max_ | AFE |
| Simulated CP-A | 397.67 | 1.37 | 251.47 | 1.25 | 3.97 | 1.85 | 389.02 | 1.48 | 69.18 | 1.40 | 0.75 | 1.00 |
| Simulated CP-B | 472.48 | 1.15 | 211.65 | 1.06 | 4.94 | 1.49 | 426.66 | 1.35 | 65.40 | 1.48 | 0.75 | 1.00 |
| Simulated CP-C | 535.03 | 1.01 | 186.91 | 1.07 | 6.04 | 1.22 | 538.19 | 1.07 | 75.07 | 1.29 | 0.75 | 1.00 |
| Simulated mean CP | 468.39 | 1.16 | 213.50 | 1.07 | 5.06 | 1.45 | 451.29 | 1.28 | 69.88 | 1.39 | 0.75 | 1.19 |

Data are presented as the mean. AFE, absolute average-fold error calculated considering observed data [1]. AUC_0-inf_, area under the plasma concentration-time curve over a dosing interval (ng·h/mL). C_max_ maximum plasma concentration (ng/mL). t_1/2_, half-life time (h). CL, clearance (mL/h/kg). t_max_, time to maximum plasma concentration (h).

**Table S2.** Individual simulated pharmacokinetic parameters of propranolol (PRO) matching individual age, Child-Pugh score, and shunt index of observed data.^2^

|  |  |  |  |  | PRO 1 mg IV | | | | | | PRO 40 mg PO | | | | | |
| --- | --- | --- | --- | --- | --- | --- | --- | --- | --- | --- | --- | --- | --- | --- | --- | --- |
| ID | Age | BMI | CP | Shunt  Index (%) | AUC_0-inf_ | | | CL | | | AUC_0-inf_ | | | C_max_ | | |
|  |  |  |  |  | Simulated | Clinical | AFE | Simulated | Clinical | AFE | Simulated | Clinical | AFE | Simulated | Clinical | AFE |
| 1 | 57 | 20.4 | B | 7 | 1832 | 889 | 2.06 | 734 | 1125 | 1.53 | 26149 | 39289 | 1.50 | 48 | 90 | 1.89 |
| 2 | 51 | 31.2 | A | 21 | 1240 | 1818 | 1.47 | 1080 | 550 | 1.96 | 20319 | 48787 | 2.40 | 50 | 59 | 1.19 |
| 5 | 47 | 21.2 | A | 12 | 1221 | 883 | 1.38 | 1108 | 1133 | 1.02 | 16339 | 22707 | 1.39 | 40 | 25 | 1.60 |
| 6 | 37 | 24.2 | A | 9 | 1217 | 791 | 1.54 | 1109 | 1265 | 1.14 | 14984 | 14143 | 1.06 | 36 | 50 | 1.39 |
| 7 | 50 | 25.4 | C | 64 | 2383 | 1752 | 1.36 | 559 | 571 | 1.02 | 71461 | 72859 | 1.02 | 109 | 63 | 1.73 |
| 8^*^ | 49 | 28.1 | B | 59 | 1792 | 7200 | 4.02 | 750 | 139 | 5.40 | 46360 | 88124 | 1.90 | 90 | 62 | 1.46 |
| 9 | 50 | 29.6 | C | 31 | 2475 | 1264 | 1.96 | 533 | 791 | 1.48 | 55098 | 49476 | 1.11 | 81 | 33 | 2.46 |
| 10 | 60 | 31.8 | A | 16 | 1330 | 1432 | 1.08 | 1020 | 698 | 1.46 | 20667 | 17894 | 1.15 | 48 | 20 | 2.41 |
| 11^*^ | 60 | 30.4 | A | 24 | 1302 | 1536 | 1.18 | 1033 | 651 | 1.59 | 21844 | 76244 | 3.49 | 53 | 132 | 2.48 |
| 12 | 51 | 20.4 | B | 43 | 1775 | 1964 | 1.11 | 754 | 606 | 1.24 | 40992 | 66527 | 1.62 | 80 | 32 | 2.51 |
| 13 | 58 | 21.3 | B | 22 | 1931 | 2540 | 1.32 | 699 | 394 | 1.77 | 35106 | 42261 | 1.20 | 63 | 53 | 1.18 |
| 14 | 67 | 26.7 | A | 13 | 1356 | 1004 | 1.35 | 1001 | 996 | 1.00 | 20577 | 44270 | 2.15 | 48 | 82 | 1.71 |
| 15^*^ | 50 | 23.5 | B | 19 | 1778 | 505 | 3.52 | 756 | 1981 | 2.62 | 30022 | 23729 | 1.27 | 56 | 59 | 1.05 |
| All |  |  |  |  | 1676 | 1778 | 1.06 | 597 | 833 | 1.40 | 34258 | 47260 | 1.38 | 60 | 65 | 1.08 |

Data are presented as the mean. ^*^outliers individuals. AFE, absolute average-fold error calculated considering observed data. Age, years. CP, Child-Pugh score. AUC_0-inf_, area under the plasma concentration-time curve over a dosing interval (ng.min/mL). C_max_ maximum plasma concentration (ng/mL). CL, clearance (mL/min).

**Table S3.** Mean simulated pharmacokinetic parameters of dexamethasone (DEX) in different liver disease conditions.

|  |  | IV | | | PO | | | | *f*_up_ |
| --- | --- | --- | --- | --- | --- | --- | --- | --- | --- |
| Dose | Population | AUC_0-24_ | CL | Vd | AUC_0-24_ | C_max_ | CL | Vd |  |
| 6 mg | Healthy | 366.61 (18) | 16.36 (14) | 87.21 (10) | 320.64 (21) | 48.30 (17) | 18.70 (13) | 100.31(13) | 0.23 |
|  | CP-A | 533.03 (17) | 11.25 (12) | 78.03 (13) | 484.15 (22) | 61.44 (19) | 12.38 (16) | 85.21 (11) | 0.25 |
|  | CP-B | 652.73 (17) | 9.18 (14) | 74.95 (12) | 590.51 (18) | 66.81 (16) | 10.15 (12) | 81.81 (11) | 0.28 |
|  | CP-C | 782.46 (14) | 7.66 (12) | 73.54 (12) | 708.24 (16) | 69.20 (15) | 8.46 (12) | 82.94 (11) | 0.34 |
|  | CP-A_shunt_ | 525.04 (17) | 11.42 (14) | 78.23 (11) | 512.23 (17) | 62.22 (15) | 11.70 (12) | 83.86 (11) | 0.25 |
|  | CP-B_shunt_ | 646.17 (16) | 9.28 (12) | 76.76 (11) | 620.46 (18) | 68.29 (16) | 9.66 (12) | 78.79 (12) | 0.28 |
|  | CP-C_shunt_ | 785.94 (17) | 7.63 (12) | 71.48 (12) | 749.54 (18) | 75.73 (18) | 8.00 (12) | 75.21 (12) | 0.34 |
| 20 mg | Healthy | 1190.15 (17) | 16.80 (12) | 90.15 (13) | 1051.33 (17) | 158.44 (15) | 19.02 (12) | 102.14 (12) | 0.23 |
|  | CP-A | 1748.29 (19) | 11.43 (14) | 78.82 (10) | 1616.10 (18) | 202.82 (17) | 12.37 (12) | 85.67 (11) | 0.25 |
|  | CP-B | 2118.08 (16) | 9.43 (12) | 77.09 (12) | 1944.64 (17) | 216.31 (14) | 10.28 (12) | 83.72 (12) | 0.28 |
|  | CP-C | 2547.98 (15) | 7.84 (12) | 72.20 (12) | 2269.89 (19) | 230.72 (15) | 8.80 (12) | 82.44 (11) | 0.34 |

Data are presented as the mean (coefficient of variation, %). AUC_0-24_, area under the plasma concentration-time curve over a dosing interval (ng.h/mL). C_max_ maximum plasma concentration (ng/mL). CL, clearance (L/h). Vd, volume of distribution (L). *f*_up_, fraction unbound in plasma. CP-A, CP-B, CP-C and CP-A_shunt_, CP-B_shunt_, CP-C_shunt_ correspond to the Child-Pugh score without and with shunting incorporated, respectively. IV and PO, intravenous and oral administration.
